# Benchmarking GPT-5 Performance and Repeatability on the Japanese National Examination for Radiological Technologists over the Past Decade (2016–2025)

**DOI:** 10.1101/2025.08.20.25333981

**Authors:** Kensuke Umehara, Junko Ota, Tatsuya Nishii, Riwa Kishimoto, Takayuki Ishida

## Abstract

**Purpose:** To evaluate GPT-5 against GPT-4o on the Japanese national examination for radiological technologists (2016–2025), assessing accuracy, repeatability, and factors influencing performance differences.

**Materials and methods:** We analyzed 1,992 multiple-choice questions spanning medical and engineering domains, including text- and image-based questions. Both models answered all questions in Japanese under identical conditions across three independent runs. Majority-vote accuracy (correct if ≥ 2 of 3 runs were correct) and first-attempt accuracy were compared using McNemar’s test. Repeatability was quantified with Fleiss’ κ. Univariable and multivariable analyses were conducted to identify question-level factors associated with GPT-5 improvements.

**Results:** GPT-5 consistently outperformed GPT-4o across all 10 exam years, achieving a majority-vote accuracy of 92.8% (95% CI: 91.5–93.8) compared with 72.4% (95% CI: 70.4–74.4) (P < .001). Repeatability was higher for GPT-5 (κ = 0.925) than GPT-4o (κ = 0.904), with correct answers in all three runs for 88.2% vs. 68.9% of items. GPT-5 achieved marked gains on text-based questions (96.5% vs. 78.1%) and substantial improvements on image-based questions (72.6% vs. 41.9%). Within medical images, significant improvements were observed for MRI, CT, and radiography, whereas performance gains were smaller for ultrasound and nuclear medicine, highlighting persistent challenges in clinically oriented image interpretation. The greatest advantages overall were observed in calculation questions (97.3% vs. 39.3%) and engineering-related domains, consistent with external benchmarks highlighting GPT-5’s strengthened reasoning.

**Conclusion:** GPT-5 demonstrated significantly higher accuracy and repeatability than GPT-4o across a decade of examinations, with especially pronounced gains in quantitative reasoning, engineering content, and diagram interpretation. Although improvements extended to medical images, performance in clinical image interpretation remained limited.

## Introduction

Large language models (LLMs) such as ChatGPT, developed by OpenAI, have recently attracted significant attention in medicine. Trained on massive datasets, these models demonstrate capabilities that surpass those of traditional natural language processing techniques. Within healthcare, a broad range of potential applications has been reported, including clinical decision support [1, 2] and medical education [3, 4].

LLM performance has been evaluated across numerous medical licensing and specialty board examinations, including the United States Medical Licensing Examination (USMLE) [5, 6], the Neurosurgery Board Exam [7], and the Neurology Board Exam [8]. In radiology, most studies have focused on board certification examinations, including the Fellowship of the Royal College of Radiologists (FRCR) [9, 10] and the European Board of Radiology (EBR) [11] in Europe, the American Board of Radiology (ABR) [10] [12] and the Canadian Royal College (CRC) [12] in North America, and the Japan Radiology Board (JRB) [13–15] in Japan. Across these settings, LLMs have demonstrated promising utility.

Radiological technologists represent another highly specialized professional group within the radiology field. They are essential healthcare providers in diagnostic imaging and radiation therapy, with formal credentialing systems established in many countries. In Japan, certification is conferred through a national examination administered by the Ministry of Health, Labour and Welfare (MHLW). The Japanese national examination for radiological technologists is notable for its breadth. In addition to testing medical knowledge related to diagnostic imaging and radiation therapy, it covers an extensive range of topics spanning the physical and engineering sciences, including medical engineering, radiation physics, radiation biology, measurement technology, safety management, medical informatics, and image processing. The examination includes both text-based questions and image-based questions, the latter incorporating medical images, graphs, and circuit diagrams. Its scope ranges from anatomy to the principles and design of multiple imaging modalities—radiography (X-ray), CT, MRI, ultrasound, and nuclear medicine—requiring proficiency in both medical and technical disciplines.

Previous studies have examined LLM performance on radiological technologist examinations, including the American Registry of Radiologic Technologists (ARRT) exam [16] and the Japanese national examination [17], but these analyses have generally been limited in question volume. Consequently, variations in performance across subject areas and by question format remain poorly characterized.

Moreover, LLMs are inherently nondeterministic, producing different outputs for the same input, and may generate hallucinations—plausible but factually incorrect responses. For any potential clinical deployment, it is therefore essential to assess reliability through repeatability testing using a wide variety of questions.

On August 7, 2025, OpenAI released GPT-5 [18]. Reported advances include substantial reductions in hallucinations, enhanced multi-step reasoning through reinforcement learning, and improved multilingual robustness, including Japanese [18]. On public benchmarks, GPT-5 demonstrated marked improvements: on competitive mathematics (AIME 2025), it achieved 94.6% (vs. 42.1% for GPT-4o) [19], and on the high-difficulty science QA benchmark GPQA, 85.7% (vs. 70.1%) [19]. Within the health domain, GPT-5 reached 67.2% on HealthBench and 46.2% on HealthBench-Hard (vs. 32.0% and 0.0% for GPT-4o, respectively) [18]. In multimodal medical reasoning, performance on MedXpertQA was 69.99% on the reasoning track and 74.37% on the understanding track, corresponding to improvements of +29.26 and +26.18 points over GPT-4o [20]. By contrast, on the radiology visual question answering dataset VQA-RAD, GPT-5 achieved 70.92%, nearly identical to GPT-4o (69.91%) [20]. Taken together, these findings suggest that improvements observed on general and non-radiology medical benchmarks may not directly translate to radiology-specific tasks, highlighting the need for domain-specific evaluation.

The purpose of this study was to evaluate GPT-5 against GPT-4o on the Japanese national examination for radiological technologists (2016–2025), quantifying accuracy, repeatability, and question-level factors associated with performance differences across all questions.

## Materials and methods

### Data collection

Multiple-choice questions from the Japanese national examination for radiological technologists administered between 2016 and 2025 were obtained from the MHLW website. Each annual examination comprises 200 questions, yielding 2,000 questions in total over the 10-year period. In accordance with official announcements, seven questions were excluded from scoring and one question was credited to all examinees; these eight questions were removed, resulting in 1,992 questions for analysis.

Each question was mapped to a subject area according to the official examination blueprint and annotated across four dimensions: question type, question style, question category, and question wording. Question type distinguished text-based questions from image-based questions, with the latter defined operationally as items that required inspection of an accompanying visual (e.g., radiologic images, diagrams, or graphs) to determine the answer. Question style indicated either the A type, in which one correct option is selected from five choices, or the X2 type, in which two correct options are selected from five choices. Question category captured the nature of the task as general (conceptual), calculation (numeric or formula-based), matching (pairing or combinations), or ordering (sequencing). Question wording was classified as either affirmative, where the question was stated affirmatively, or negative, which included questions such as “Which of the following is incorrect?” requiring identification of an exception. Image category was defined as follows: when the attached visual was a medical image, it was classified by imaging modality (e.g., MRI, X-ray (radiography), CT, ultrasound, nuclear medicine, fundus); when the visual was non-medical, it was classified by image type (e.g., graphs/plots, circuit diagrams, schematic diagrams); items not assignable to either were labeled “Others”. Text length was measured as character count and categorized into quartiles: Very short (< Q1), Short (Q1 to < Median), Medium (Median to < Q3), and Long (≥ Q3).

All exam materials were originally provided as PDF files. Questions and official answer keys were extracted using Adobe Acrobat Pro (Adobe Inc., San Jose, CA, USA). Text content was exported as plain text, and images accompanying image-based questions were exported directly as 300 dpi PNG files.

### Study design

This retrospective, paired study compared two LLMs (GPT-5 and GPT-4o) using publicly available examination materials from the 2016–2025 Japanese national examination for radiological technologists. Because this study used only publicly available exam questions and involved no human participants or animal subjects, institutional review board approval and informed consent were not required.

After prespecified exclusions, both models (GPT-5 and GPT-4o, OpenAI) answered the same 1,992 questions under identical input formatting with automated exact-match scoring. Each question was evaluated in three independent runs per model. All runs were executed between August 8 and August 13, 2025. Model inferences were performed via API with fixed, model-specific generation settings held constant across the study: for GPT-5, reasoning_effort = “medium” and verbosity = “low”; for GPT-4o, temperature = 0 and max_tokens = 10. For every item, we used the following prompt, with images attached when applicable: “You will be given a question from the Japanese national examination for radiological technologists. Use domain knowledge in radiation (medicine, physics, biology, diagnostic imaging, and relevant regulations) to answer. For the following question, provide only the number of the single best option (1–5). If the question explicitly instructs you to select two options, provide exactly two options. Do not output any reasons, explanations, calculation steps, or reasoning processes.” The question stem and answer options were entered directly into the models in their original Japanese, exactly as published. When present, accompanying images were attached in the original order as 300 dpi PNGs. All API calls were executed using Python 3.11.5 with the OpenAI Python API library (version 1.99.3).

The primary outcome was overall accuracy, defined as majority-vote accuracy (correct if at least two of three runs were correct). The secondary outcome was first-attempt accuracy (the first of the three runs). Both endpoints were prespecified to capture complementary use cases: first-attempt accuracy reflects single-shot, exam-like conditions, whereas majority-vote accuracy reduces sampling variance from decoding randomness and provides a reliability-adjusted estimate obtainable with minimal re-queries. Prespecified subgroup analyses were conducted by question type, question style, question category, question wording, subject area, image category, and text-length quartiles.

### Statistical analysis

Accuracy was computed at the item level for each model using two prespecified metrics: first-attempt accuracy (the first of three independent runs) and majority-vote accuracy (defined as correct if at least two of three runs were correct). Between-model comparisons were performed using McNemar’s test on paired majority-vote outcomes. For repeatability, agreement across the three runs per model was quantified using Fleiss’ kappa.

To identify attributes associated with questions answered correctly by GPT-5 but incorrectly by GPT-4o, we first performed univariable Fisher’s exact tests for each candidate attribute, with multiplicity controlled for using the Benjamini–Hochberg false discovery rate (FDR). We then fitted a multivariable logistic regression model with GPT-5 improvement as the dependent variable. Candidate predictors included question type, question style, question category, question wording, text length, and subject area. Variable selection was performed via stepwise selection based on Akaike’s information criterion (AIC). All analyses were conducted in R (version 4.4.3; R Foundation for Statistical Computing, Vienna, Austria). A p-value < 0.05 was considered statistically significant.

## Results

### Question characteristics

Figure 1 shows the distribution of text-based and image-based questions across subject areas and question characteristics. Of the 1,992 questions, 84.4% were text-based, whereas 15.6% required image interpretation. The proportion of image-based questions exceeded half only in Medical Engineering (54.4%) and approached half in X-ray Imaging Technology (47.5%) and Diagnostic Imaging (47.0%). It was also relatively high in Image Engineering (39.2%). In contrast, some domains contained no image-based questions—Radiation Biology, Radiochemistry, Radiation Safety Management, and Patient Safety Management. Others were almost entirely text-based, including Basic Medical Sciences (1.0% image-based), Nuclear Medicine Technology (3.6%), Radiation Measurement (6.0%), Medical Imaging Technology (6.0%), Radiation Therapy Technology (6.1%), Radiation Physics (9.0%), and Medical Imaging Informatics (15.0%).

**Figure 1.**
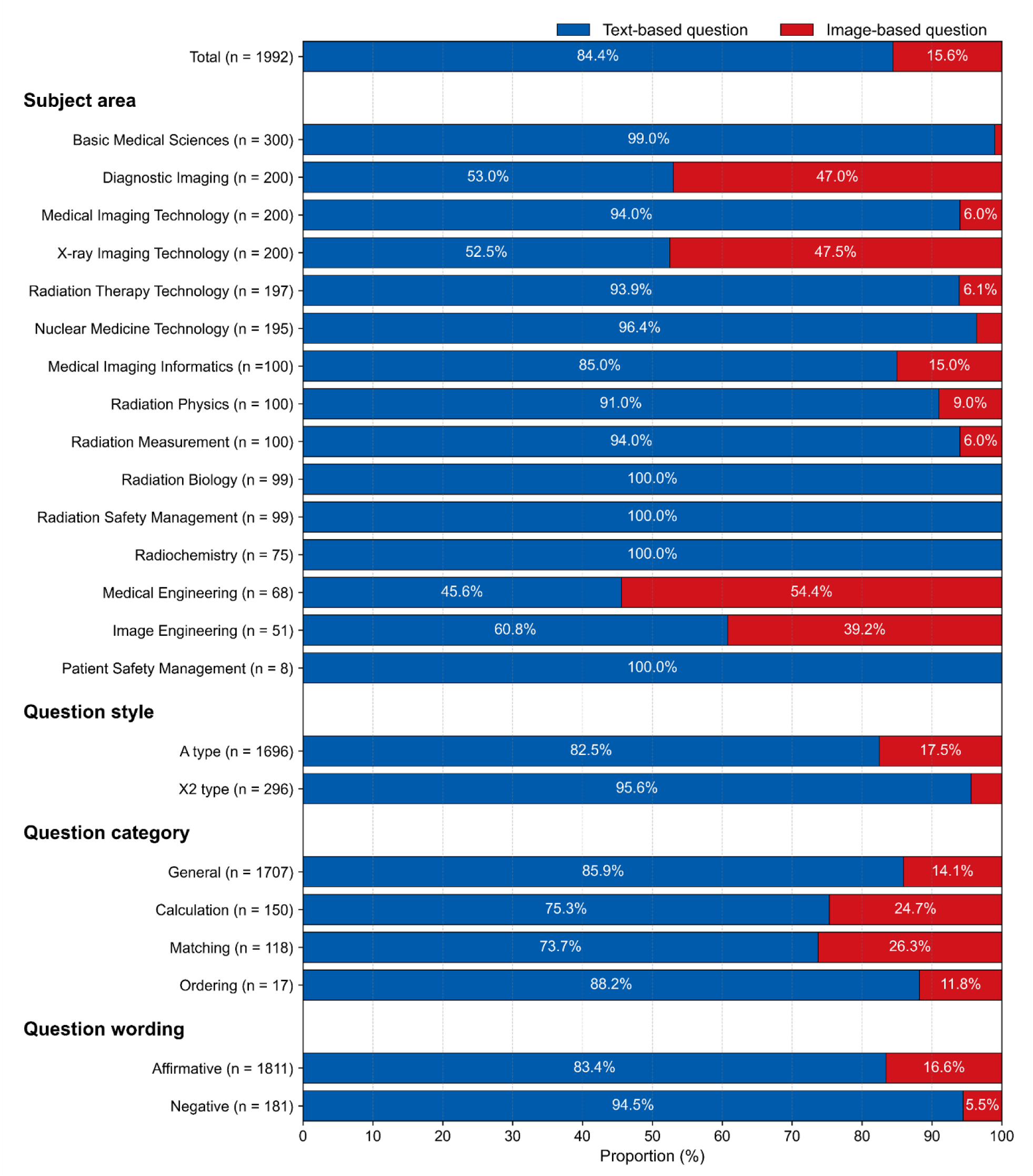
Distribution of text-based and image-based questions in the Japanese national examination for radiological technologists (2016–2025), according to subject area, question style, question category, and question wording.

Question style and category showed similar heterogeneity. X2-type questions were overwhelmingly text-based (95.6% text vs. 4.4% image), whereas A-type questions included a larger image component (82.5% text vs. 17.5% image). By category, images were most frequent in matching and calculation questions (26.3% and 24.7% image-based, respectively), followed by general (14.1%) and ordering (11.8%). For question wording, affirmative questions more often incorporated images than negative questions (16.6% vs. 5.5%). Across the 1,992 questions, the median text length of the question was 86 characters (IQR: 53–123).

### Overall accuracy

Table 1 summarizes overall and year-wise accuracy across 1,992 questions. Pooled majority-vote accuracy was 92.8% (95% CI: 91.5–93.8) for GPT-5, compared with 72.4% (95% CI: 70.4–74.4) for GPT-4o—an absolute difference of 20.4 percentage points (pp). On first attempts, accuracy was 91.8% (95% CI: 90.5– 92.9) for GPT-5, compared with 72.8% (95% CI: 70.8–74.7) for GPT-4o (difference, 19.0 pp). By McNemar’s test on paired majority-vote outcomes, the between-model difference was statistically significant overall and in every exam year (all P < .001). Year-by-year majority-vote gaps ranged from 16.0 to 24.7 pp, and first-attempt gaps from 13.5 to 22.8 pp, with GPT-5 consistently outperforming GPT- 4o. Both models exceeded Japan’s passing threshold for the national examination for radiological technologists (≥60% of total points) in every year.

**Table 1.** Overall and year-wise accuracy of GPT-5 and GPT-4o on the Japanese national examination for radiological technologists (2016–2025).

| Year | n | Majority-vote accuracy |  |  |  | First-attempt accuracy |  |  |  |
| --- | --- | --- | --- | --- | --- | --- | --- | --- | --- |
| | | GPT-5 | GPT-4o | $\Delta$ (GPT-5 | | GPT-5 | GPT-4o | $\Delta$ (GPT-5 | |
|  |  |  |  | - GPT-4o) | *P* value |  |  | - GPT-4o) |  |
| Total | 1992 | 92.8% [91.5–93.8] | 72.4% [70.4–74.4] | 20.4 | <.001 | 91.8% [90.5–92.9] | 72.8% [70.8–74.7] | 19.0 |  |
| 2025 | 200 | 92.0% [87.4–95.0] | 74.5% [68.0–80.0] | 17.5 | <.001 | 91.5% [86.8–94.6] | 73.5% [67.0–79.1] | 18.0 |  |
| 2024 | 200 | 93.0% [88.6–95.8] | 72.0% [65.4–77.8] | 21.0 | <.001 | 92.0% [87.4–95.0] | 71.5% [64.9–77.3] | 20.5 |  |
| 2023 | 200 | 91.5% [86.8–94.6] | 72.0% [65.4–77.8] | 19.5 | <.001 | 91.0% [86.2–94.2] | 73.0% [66.5–78.7] | 18.0 |  |
| 2022 | 197 | 92.4% [87.8–95.3] | 72.1% [65.4–77.9] | 20.3 | <.001 | 92.9% [88.4–95.7] | 70.1% [63.3–76.0] | 22.8 |  |
| 2021 | 199 | 95.0% [91.0–97.2] | 73.4% [66.8–79.0] | 21.6 | <.001 | 93.5% [89.1–96.1] | 74.4% [67.9–79.9] | 19.1 |  |
| 2020 | 197 | 91.9% [87.2–94.9] | 75.6% [69.2–81.1] | 16.3 | <.001 | 91.9% [87.2–94.9] | 76.6% [70.3–82.0] | 15.3 |  |
| 2019 | 200 | 93.5% [89.2–96.2] | 77.5% [71.2–82.7] | 16.0 | <.001 | 91.5% [86.8–94.6] | 78.0% [71.8–83.2] | 13.5 |  |
| 2018 | 200 | 93.5% [89.2–96.2] | 69.5% [62.8–75.5] | 24.0 | <.001 | 91.0% [86.2–94.2] | 70.0% [63.3–75.9] | 21.0 |  |
| 2017 | 199 | 91.0% [86.2–94.2] | 66.3% [59.5–72.5] | 24.7 | <.001 | 91.0% [86.2–94.2] | 68.8% [62.1–74.9] | 22.2 |  |
| 2016 | 200 | 94.0% [89.8–96.5] | 71.5% [64.9–77.3] | 22.5 | <.001 | 92.0% [87.4–95.0] | 72.5% [65.9–78.2] | 19.5 |  |

Square brackets indicate 95% confidence intervals [CI].
P values are from McNemar’s test comparing majority-vote accuracy between GPT-5 and GPT-4o
Majority-vote = counted correct when two or more of the three trials are correct, First-attempt = counted correct when the first of the three trials is correct, pp = percentage points.

Table 2 shows that GPT-5 outperformed GPT-4o across all strata. By question type, accuracy was higher for text than image items, with clear advantages for GPT-5 in both: text, 96.5% (95% CI: 95.5–97.3) vs. 78.1% (95% CI: 76.0–80.0); image, 72.6% (95% CI: 67.4–77.2) vs. 41.9% (95% CI: 36.6–47.5) (both P < .001). By style, GPT-5 exceeded GPT-4o for A-type (93.2% [95% CI: 91.9–94.3] vs. 72.9% [95% CI: 70.7–74.9]) and X2-type questions (90.5% [95% CI: 86.7–93.4] vs. 69.9% [95% CI: 64.5–74.9]) (both P < .001). Across categories, the sharpest contrast was for calculation (97.3% [95% CI: 93.3–99.0] vs. 39.3% [95% CI: 31.9–47.3], P < .001); matching (83.9% [95% CI: 76.2–89.4] vs. 57.6% [95% CI: 48.6–66.2], P < .001) and general (93.0% [95% CI: 91.7–94.1] vs. 76.3% [95% CI: 74.3–78.3], P < .001) also favored GPT-5, whereas ordering (94.1% [95% CI: 73.0–99.0] vs. 76.5% [95% CI: 52.7–90.4]) was not significant (P = .248). For wording, GPT-5 led for both affirmative (92.5% [95% CI: 91.2–93.7] vs. 72.0% [95% CI: 69.9–74.0], P < .001) and negative questions (95.0% [95% CI: 90.8–97.4] vs. 76.8% [95% CI: 70.1–82.3], P < .001). By subject area, advantages were broad and most pronounced in Medical Engineering (95.6% [95% CI: 87.8–98.5] vs. 42.6% [95% CI: 31.6–54.5], P < .001) and Image Engineering (92.2% [95% CI: 81.5–96.9] vs. 51.0% [95% CI: 37.7–64.1], P < .001); even in Basic Medical Sciences, where both were near ceiling, GPT-5 remained higher (99.0% [95% CI: 97.1–99.7] vs. 96.0% [95% CI: 93.1–97.7], P = .008). First-attempt analyses paralleled these patterns.

**Table 2.**
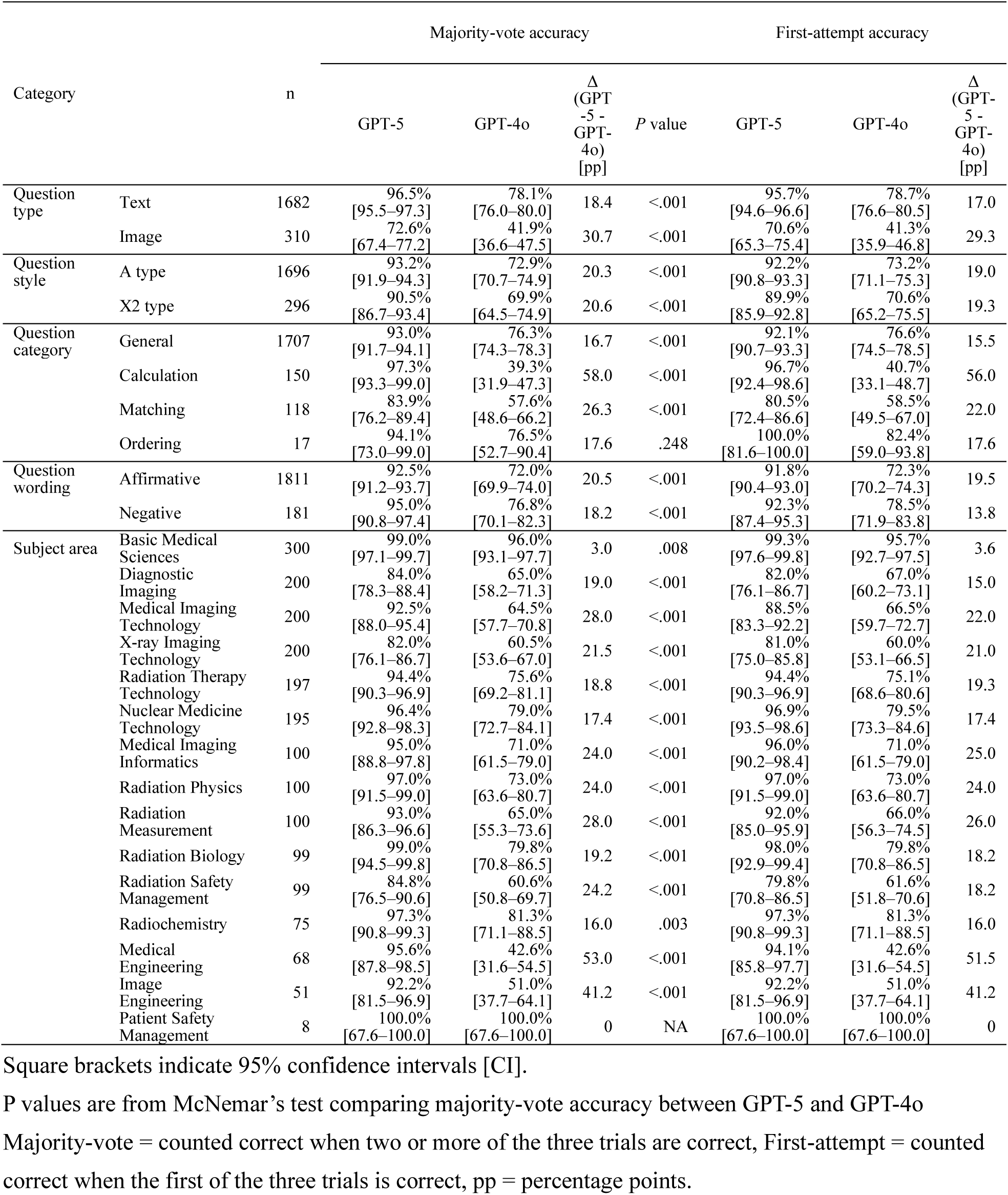
Accuracy of GPT-5 and GPT-4o by question type, style, category, wording, and subject area on the Japanese national examination for radiological technologists.

Table 3 shows that GPT-5 also outperformed GPT-4o across all image categories on the majority-vote accuracy with statistical significance in several high-volume categories. Among medical images, GPT-5 achieved higher performance for MRI (67.2% [95% CI: 54.7–77.7] vs. 52.5% [95% CI: 40.2–64.5], P = .033), radiography (X-ray) (64.3% [95% CI: 51.2–75.5] vs. 41.1% [95% CI: 29.2–54.1], P = .006), and CT (70.6% [95% CI: 53.8–83.2] vs. 38.2% [95% CI: 23.9–55.0], P = .015). Differences for ultrasound (64.3% [95% CI: 45.8–79.3] vs. 32.1% [95% CI: 17.9–50.7], P = .052) did not reach significance, and the very small categories nuclear medicine (n = 7; 85.7% [95% CI: 48.7–97.4] vs. 14.3% [95% CI: 2.6–51.3], P = .074) and fundus (n = 4; 75.0% [95% CI: 30.1–95.4] vs. 50.0% [95% CI: 15.0–85.0], P = 1.0) were also not significant. Among non-medical images, GPT-5 showed clear advantages for graphs/plots (78.7% [95% CI: 65.1–88.0] vs. 38.3% [95% CI: 25.8–52.6], P < .001) and circuit diagrams (94.3% [95% CI: 81.4–98.4] vs. 42.9% [95% CI: 28.0–59.1] , P < .001), whereas schematic diagrams (P = .221) and other images (P = .096) did not reach statistical significance.

**Table 3.** Accuracy of GPT-5 and GPT-4o by image category on the Japanese national examination for radiological technologists (2016–2025).

| Image category |  | n | Majority-vote accuracy |  |  |  | First-attempt accuracy |  |  |
| --- | --- | --- | --- | --- | --- | --- | --- | --- | --- |
| | | | GPT-5 | GPT-4o | $\Delta$ (GPT-5 - GPT-4o) [pp] | *P* value | GPT-5 | GPT-4o | $\Delta$ (GPT-5 - GPT-4o) [pp] |
| Medical images | MRI | 61 | 67.2%<br>[54.7–77.7] | 52.5%<br>[40.2–64.5] | 14.7 | .033 | 62.3%<br>[49.7–73.4] | 50.8%<br>[38.6–62.9] | 11.5 |
|  | X-ray | 56 | 64.3%<br>[51.2–75.5] | 41.1%<br>[29.2–54.1] | 23.2 | .006 | 64.3%<br>[51.2–75.5] | 37.5%<br>[26.0–50.6] | 26.8 |
|  | CT | 34 | 70.6%<br>[53.8–83.2] | 38.2%<br>[23.9–55.0] | 32.4 | .015 | 70.6%<br>[53.8–83.2] | 38.2%<br>[23.9–55.0] | 32.4 |
|  | Ultrasound | 28 | 64.3%<br>[45.8–79.3] | 32.1%<br>[17.9–50.7] | 32.2 | .052 | 60.7%<br>[42.4–76.4] | 35.7%<br>[20.7–54.2] | 25.0 |
|  | Nuclear medicine | 7 | 85.7%<br>[48.7–97.4] | 14.3% [2.6–51.3] | 71.4 | .074 | 85.7%<br>[48.7–97.4] | 28.6%<br>[8.2–64.1] | 57.1 |
|  | Fundus | 4 | 75.0%<br>[30.1–95.4] | 50.0%<br>[15.0–85.0] | 25.0 | 1.0 | 75.0%<br>[30.1–95.4] | 50.0%<br>[15.0–85.0] | 25.0 |
| Non-medical images | Graphs/Plots | 47 | 78.7%<br>[65.1–88.0] | 38.3%<br>[25.8–52.6] | 40.4 | <.001 | 74.5%<br>[60.5–84.7] | 36.2%<br>[24.0–50.5] | 38.3 |
|  | Circuit diagrams | 35 | 94.3%<br>[81.4–98.4] | 42.9%<br>[28.0–59.1] | 51.4 | <.001 | 94.3%<br>[81.4–98.4] | 42.9%<br>[28.0–59.1] | 51.4 |
|  | Schematic diagrams | 14 | 64.3%<br>[38.8–83.7] | 35.7%<br>[16.3–61.2] | 28.6 | .221 | 64.3%<br>[38.8–83.7] | 35.7%<br>[16.3–61.2] | 28.6 |
|  | Others | 32 | 75.0%<br>[57.9–86.7] | 53.1%<br>[36.4–69.1] | 21.9 | .096 | 75.0%<br>[57.9–86.7] | 53.1%<br>[36.4–69.1] | 21.9 |

Square brackets indicate 95% confidence intervals [CI].
P values are from McNemar’s test comparing majority-vote accuracy between GPT-5 and GPT-4o
Majority-vote = counted correct when two or more of the three trials are correct, First-attempt = counted correct when the first of the three trials is correct, pp = percentage points.

### Repeatability analysis

Fleiss’ kappa across the three runs indicated high repeatability for both models: GPT-5, 0.925 (95% CI: 0.915–0.935); GPT-4o, 0.904 (95% CI: 0.894–0.914). Across all questions, the proportion answered correctly in all three runs was 88.2% (1756/1992) for GPT-5, compared with 68.9% (1373/1992) for GPT- 4o, whereas consistently incorrect responses on all three runs were 2.8% (55/1992) compared with 19.1% (381/1992). Among text questions, the proportion answered correctly on all three runs was 93.3% (1569/1682) for GPT-5, compared with 75.3% (1266/1682) for GPT-4o. Among image-based questions, the corresponding proportions were 60.3% (187/310) and 34.5% (107/310), respectively. A full stratified breakdown is provided in Supplemental Table S1.

### Associations with GPT-5 improvements

To identify the types of questions where GPT-5 achieved gains over GPT-4o, we compared items that were answered correctly by GPT-5 but incorrectly by GPT-4o. Of the 1,992 questions, 441 (22.1%) met this criterion. Figure 2 shows the univariable associations across question characteristics, and Figure 3 shows those by subject area; a volcano plot summarizing all features appears in Supplemental Figure S1. Factors more commonly associated with GPT-5 improvements included image-based questions (OR = 2.59, 95% CI: 2.00–3.35, FDR-adjusted P < .001) and calculation questions (OR = 6.15, 95% CI: 4.36–8.69, FDR-adjusted P < .001). Additional associations were observed for long text (OR = 1.46, 95% CI: 1.15–1.84, FDR-adjusted P = .007); for medical image types—ultrasound (OR = 3.12, 95% CI: 1.49–6.53, FDR-adjusted P = .018), nuclear medicine (OR = 7.81, 95% CI: 1.74–34.96, FDR-adjusted P = .033), and CT (OR = 2.53, 95% CI: 1.28–5.01, FDR-adjusted P = .0369); and for non-medical image categories, including circuit diagrams (OR = 4.82, 95% CI: 2.47–9.41, FDR-adjusted P < .001) and graphs/plots (OR = 2.94, 95% CI: 1.65–5.25, FDR-adjusted P = .003). Among subject areas, Medical Engineering (OR = 4.76, 95% CI: 2.93–7.75, FDR-adjusted P < .001) and Image Engineering (OR = 3.00, 95% CI: 1.72–5.24, FDR-adjusted P = .002), and Medical Imaging Technology (OR = 1.51, 95% CI: 1.09–2.09, FDR-adjusted P = .037) were associated with GPT-5 improvements.

**Figure 2.**
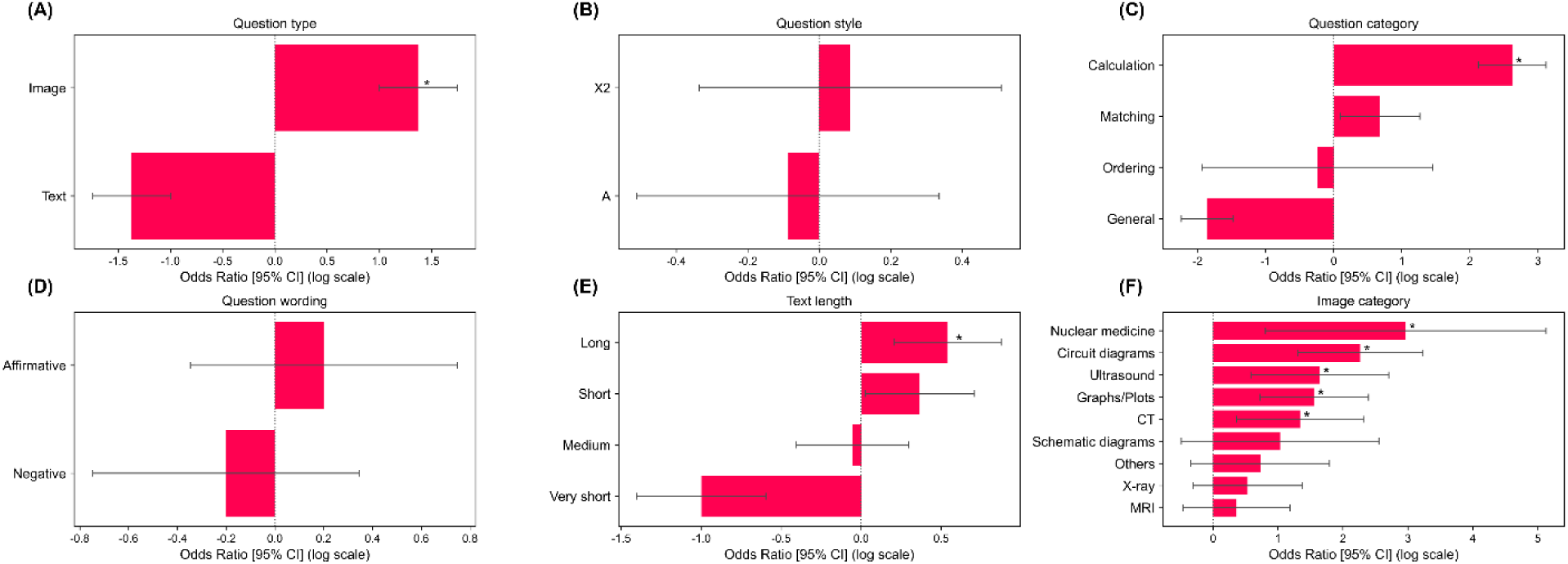
Univariable odds ratios (95% CI) for factors associated with items answered correctly by GPT-5 but incorrectly by GPT-4o. Panels A–F show results by (A) question type, (B) question style, (C) question category, (D) question wording, (E) text length, and (F) image category. CI = confidence interval.

**Figure 3.**
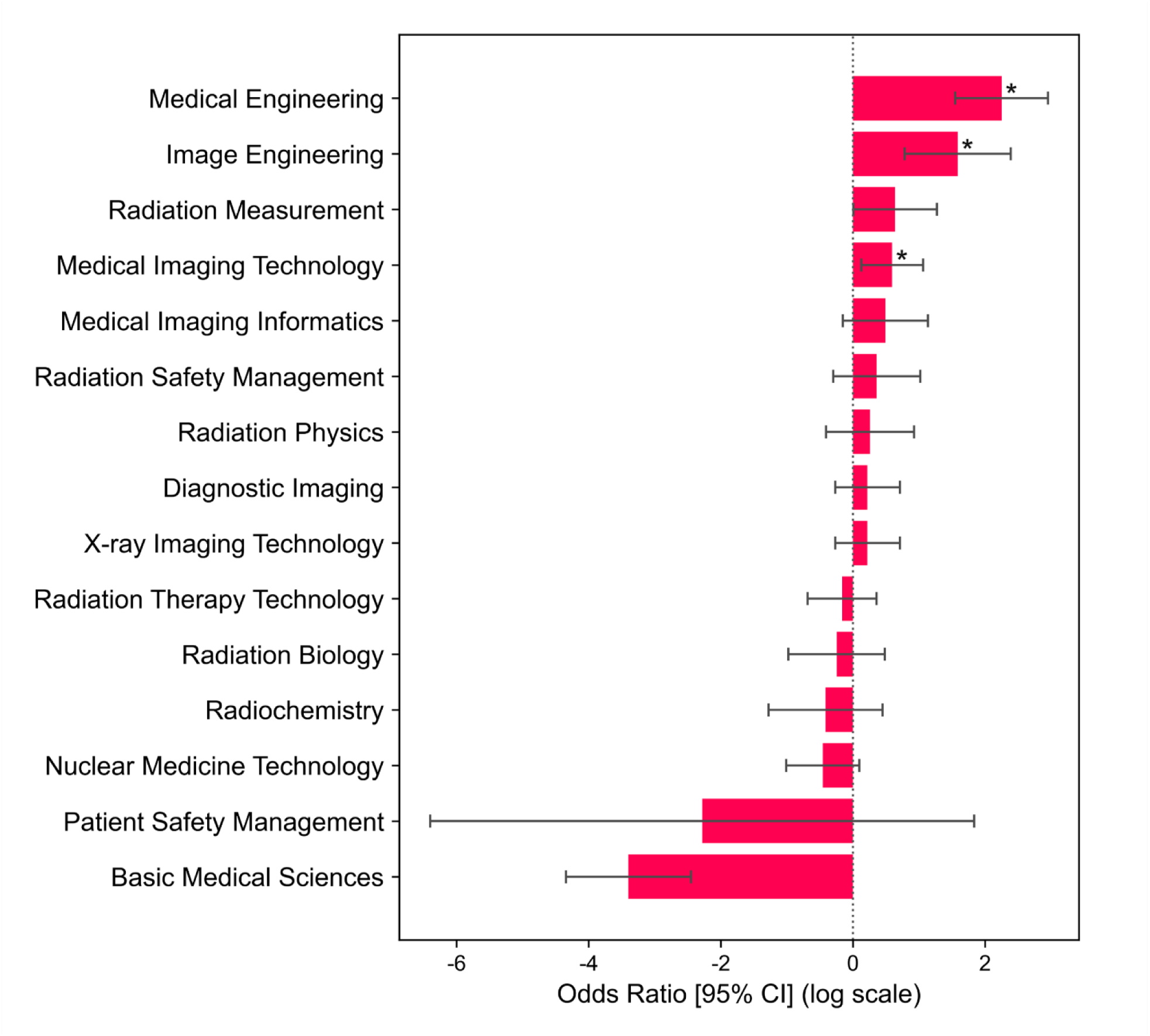
Univariable odds ratios (95% CI) for subject areas associated with items answered correctly by GPT-5 but incorrectly by GPT-4o. CI = confidence interval.

An AIC-based stepwise multivariable logistic regression retained question type, question style, question category, and subject area, whereas question wording and text length were not selected. Figure 4 shows adjusted odds ratios (aORs) from this model. Independent associations with GPT-5 improvements were observed for calculation questions (aOR = 4.53, 95% CI: 3.05–6.73, FDR-adjusted P < .001), image-based questions (aOR = 2.11, 95% CI: 1.52–2.94, FDR-adjusted P < .001), and X2-style questions (aOR = 1.39, 95% CI: 1.01–1.92, FDR-adjusted P = .0498). Multiple subject areas remained significant, led by Image Engineering (aOR = 13.91, 95% CI: 5.66–34.20, FDR-adjusted P < .001) and Medical Engineering (aOR = 12.19, 95% CI: 5.03–29.58, FDR-adjusted P < .001). Other significant areas showed aORs from 5.81 to 10.68 (all FDR-adjusted P < .001).

**Figure 4.**
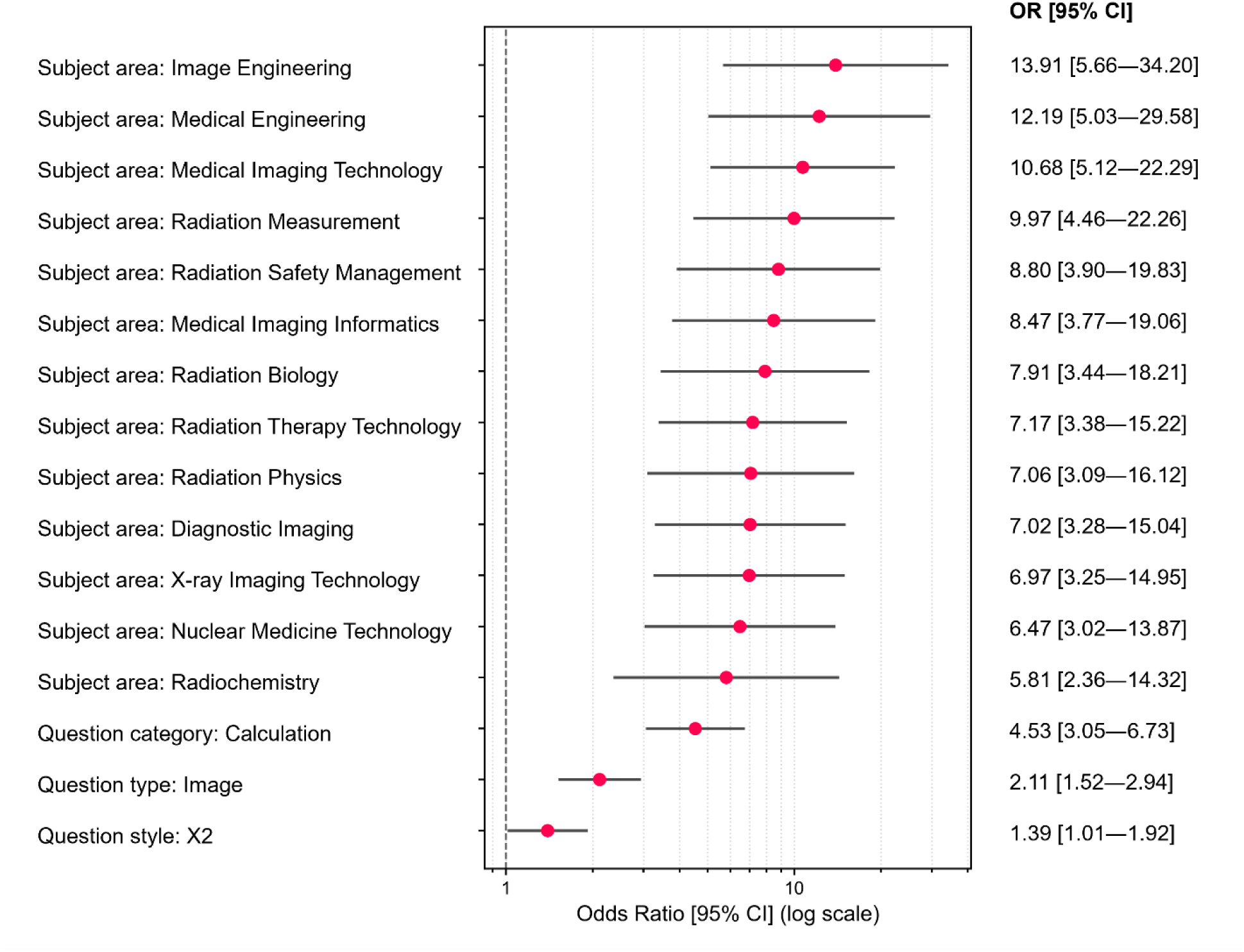
Multivariable logistic regression showing adjusted odds ratios (95% CI) for factors independently associated with items answered correctly by GPT-5 but incorrectly by GPT-4o. CI = confidence interval, OR = odds ratio.

## Discussion

The present study evaluated state-of-the-art GPT-5 against GPT-4o using the Japanese national examination for radiological technologists, encompassing 1,992 questions administered over the past decade. This examination integrates both medical and engineering aspects of radiological sciences and includes a diverse range of text-based and image-based questions. To ensure a nuanced assessment, we conducted stratified analyses across multiple dimensions, including question type, style, category, wording, subject area, image category, and text length. Model performance was further examined through a three-trial design with majority voting, enabling simultaneous assessment of repeatability and practical stability. Across all questions, GPT-5 consistently outperformed GPT-4o, achieving a majority-vote accuracy of 92.8% (95% CI: 91.5–93.8) compared with 72.4% (95% CI: 70.4–74.4), with significant differences confirmed by McNemar’s testing in every examination year (all P < .001) (Table 1).

Repeatability was also superior for GPT-5, with Fleiss’ κ of 0.925 compared with 0.904 for GPT-4o. These results demonstrate that GPT-5 provides both higher accuracy and more stable performance than GPT-4o under exam-like conditions.

GPT-5 also demonstrated clear improvements over GPT-4o for image-based questions, although absolute performance remained lower than for text-based items. Majority-vote accuracy reached 96.5% (95% CI: 95.5–97.3) for text-based questions compared with 72.6% (95% CI: 67.4–77.2) for image-based questions. Within the image domain, significant gains were observed for MRI, radiography, and CT. By contrast, performance was lower and did not reach statistical significance for ultrasound, nuclear medicine, and fundus imaging, likely reflecting the limited number of items in these categories. As shown in Table 3, accuracy for medical images as a group was consistently lower than for non-medical images such as graphs/plots (78.7%) and circuit diagrams (94.3%), suggesting that GPT-5 still faces greater challenges in clinically oriented image interpretation than in schematic or diagrammatic tasks. These findings indicate that while GPT-5 achieved substantial gains in image interpretation compared with GPT-4o, its performance remained inferior to text-based accuracy and showed variability across imaging modalities.

To elucidate the factors underlying GPT-5’s performance gains, we conducted both univariable and multivariable analyses. In univariable models, longer question length was positively associated with accuracy, but text length and question wording (affirmative or negative) were excluded from the final multivariable model, suggesting that these factors were not independent determinants of performance and may be confounded by question category or subject area. By contrast, multivariable models confirmed that calculation questions, image-based questions, X2-type questions, and multiple subject areas—including Image Engineering and Medical Engineering—remained independently associated with higher GPT-5 accuracy. The independent effect of X2-type questions suggests that GPT-5 has developed greater robustness in reasoning through multiple-response options. Moreover, the particularly large gains in calculation and engineering-related questions imply strengthened abilities in mathematical reasoning and technical diagram interpretation. These trends are consistent with GPT-5’s advances reported on external benchmarks such as AIME, GPQA, and MedXpertQA, which emphasize mathematical and multimodal reasoning [19]. Notably, whereas a recent study (e.g., VQA-RAD) has reported relatively modest improvements in pure medical image interpretation [20], our findings suggest that enhanced understanding of diagrams, schematics, and physics- or engineering-related content may represent a key driver of GPT- 5’s advantage.

As noted in OpenAI’s system card [18], our findings confirm that GPT-5 demonstrates enhanced robustness in Japanese, which likely contributed to its improved overall performance. Although challenges remain in medical image interpretation, with some modalities such as ultrasound showing relatively lower accuracy, these limitations appear less pronounced outside image-focused tasks. From an educational standpoint, GPT-5 shows considerable promise as a supplementary tool, particularly for domains involving calculations, engineering content, and schematic or diagrammatic reasoning. Potential applications include generating worked examples for numerical problems, extracting key elements from diagrams, and developing practice items for examination preparation. However, for tasks that involve direct clinical image interpretation or have implications for patient safety, human oversight remains essential.

This study has several limitations that should be acknowledged. First, the analysis was conducted exclusively in Japanese, and generalizability to other languages such as English remains untested. Second, performance was assessed on the Japanese national examination for radiological technologists and therefore may not directly translate to free-text or unstructured data encountered in clinical practice. Third, image inputs were standardized as 300 dpi PNGs, and it remains unclear whether changes in resolution or format would influence model performance. Fourth, statistical power was limited for certain small categories, such as nuclear medicine (n = 7) and fundus imaging (n = 4), which did not reach significance; expanding the sample size in these domains will be necessary for more definitive evaluation. The relatively modest gains for medical images may also reflect the smaller representation of such data in training corpora compared with non-medical images. Fifth, because the examination questions are publicly available, the possibility of training data leakage cannot be entirely excluded. Sixth, our evaluation focused on answer accuracy; further work is needed to assess the validity of the model’s reasoning processes. Finally, safety considerations remain critical, and systematic analysis of error patterns—particularly items answered incorrectly in all three trials—will be essential to identify potential risks. Future research should also examine the appropriateness of GPT-5 as an educational tool by evaluating the validity of its explanations and outputs through expert review.

In conclusion, we demonstrated that GPT-5 consistently outperformed GPT-4o with markedly higher accuracy and repeatability by evaluating a decade of the Japanese national examination for radiological technologists, which spans both medical and engineering domains. The most pronounced improvements were observed in calculation, engineering, and diagram-based tasks, reflecting strengthened reasoning and technical understanding. Although GPT-5 also showed clear gains in image-based questions compared with GPT-4o, performance in medical image interpretation remained relatively limited. These findings highlight the potential utility of GPT-5 as an educational aid, particularly in domains involving quantitative reasoning and technical content. At the same time, its outputs should not be relied upon for final image interpretation or decisions with direct implications for patient safety, where human oversight remains indispensable.

## Data Availability

The datasets generated or analyzed during the study are available from the corresponding author on reasonable request.

## Acknowledgements

During the preparation of this work, the first author used ChatGPT (OpenAI, GPT-5) to improve the readability and language of the manuscript. After using ChatGPT, all outputs were carefully reviewed, revised, and edited by the first author, and all authors assume full responsibility for the final content.

## Ethical statement

This study used only publicly available examination questions and did not involve human participants or animals. Therefore, institutional review board approval and informed consent were not required.

## References

[1] S. Liu, A.P. Wright, B.L. Patterson, J.P. Wanderer, R.W. Turer, S.D. Nelson, A.B. McCoy, D.F. Sittig, A. Wright, Using AI-generated suggestions from ChatGPT to optimize clinical decision support, Journal of the American Medical Informatics Association. 30 (7) (2023). 1237–1245. 10.1093/jamia/ocad072.

[2] J. Ferdush, M. Begum, S.T. Hossain, ChatGPT and Clinical Decision Support: Scope, Application, and Limitations, Ann Biomed Eng. 52 (5) (2024). 1119–1124. 10.1007/s10439-023-03329-4.

[3] H. Ba, L. Zhang, Z. Yi, Enhancing clinical skills in pediatric trainees: a comparative study of ChatGPT-assisted and traditional teaching methods, BMC Med Educ. 24 (1) (2024). 558. 10.1186/s12909-024-05565-1.

[4] Z. Hui, Z. Zewu, H. Jiao, C. Yu, Application of ChatGPT-assisted problem-based learning teaching method in clinical medical education, BMC Med Educ. 25 (1) (2025). 50. 10.1186/s12909-024-06321-1.

[5] T.H. Kung, M. Cheatham, A. Medenilla, C. Sillos, L. De Leon, C. Elepano, M. Madriaga, R. Aggabao, G. Diaz-Candido, J. Maningo, V. Tseng, Performance of ChatGPT on USMLE: Potential for AI-assisted medical education using large language models, PLOS Digit Health. 2 (2) (2023). e0000198. 10.1371/journal.pdig.0000198.

[6] V. Lievin, C.E. Hother, A.G. Motzfeldt, O. Winther, Can large language models reason about medical questions?, Patterns (N Y). 5 (3) (2024). 100943. 10.1016/j.patter.2024.100943.

[7] R. Ali, O.Y. Tang, I.D. Connolly, P.L. Zadnik Sullivan, J.H. Shin, J.S. Fridley, W.F. Asaad, D. Cielo, A.A. Oyelese, C.E. Doberstein, Z.L. Gokaslan, A.E. Telfeian, Performance of ChatGPT and GPT-4 on Neurosurgery Written Board Examinations, Neurosurgery. 93 (6) (2023). 1353–1365. 10.1227/neu.0000000000002632.

[8] T.C. Chen, E. Multala, P. Kearns, J. Delashaw, A. Dumont, D. Maraganore, A. Wang, Assessment of ChatGPT’s performance on neurology written board examination questions, BMJ Neurol Open. 5 (2) (2023). e000530. 10.1136/bmjno-2023-000530.

[9] S. Ariyaratne, N. Jenko, A. Mark Davies, K.P. Iyengar, R. Botchu, Could ChatGPT Pass the UK Radiology Fellowship Examinations?, Acad Radiol. 31 (5) (2024). 2178–2182. 10.1016/j.acra.2023.11.026.

[10] A. Sood, N. Mansoor, C. Memmi, M. Lynch, J. Lynch, Generative pretrained transformer-4, an artificial intelligence text predictive model, has a high capability for passing novel written radiology exam questions, Int J Comput Assist Radiol Surg. 19 (4) (2024). 645–653. 10.1007/s11548-024-03071-9.

[11] M.S. Besler, L. Oleaga, V. Junquero, C. Merino, Evaluating GPT-4o’s Performance in the Official European Board of Radiology Exam: A Comprehensive Assessment, Acad Radiol. 31 (11) (2024). 4365–4371. 10.1016/j.acra.2024.09.005.

[12] R. Bhayana, S. Krishna, R.R. Bleakney, Performance of ChatGPT on a Radiology Board-style Examination: Insights into Current Strengths and Limitations, Radiology. 307 (5) (2023). e230582. 10.1148/radiol.230582.

[13] Y. Toyama, A. Harigai, M. Abe, M. Nagano, M. Kawabata, Y. Seki, K. Takase, Performance evaluation of ChatGPT, GPT-4, and Bard on the official board examination of the Japan Radiology Society, Jpn J Radiol. 42 (2) (2024). 201–207. 10.1007/s11604-023-01491-2.

[14] T. Nakaura, N. Yoshida, N. Kobayashi, Y. Nagayama, H. Uetani, M. Kidoh, S. Oda, Y. Funama, T. Hirai, Performance of Multimodal Large Language Models in Japanese Diagnostic Radiology Board Examinations (2021-2023), Acad Radiol. 32 (5) (2025). 2394–2401. 10.1016/j.acra.2024.10.035.

[15] T. Nakaura, H. Takamure, N. Kobayashi, K. Shiraishi, N. Yoshida, Y. Nagayama, H. Uetani, M. Kidoh, Y. Funama, T. Hirai, Evaluating the Performance of Reasoning Large Language Models on Japanese Radiology Board Examination Questions, Acad Radiol. 32 (8) (2025). 4347–4354. 10.1016/j.acra.2025.04.060.

[16] Y. Al-Naser, F. Halka, B. Ng, D. Mountford, S. Sharma, K. Niure, C. Yong-Hing, F. Khosa, C. Van der Pol, Evaluating Artificial Intelligence Competency in Education: Performance of ChatGPT-4 in the American Registry of Radiologic Technologists (ARRT) Radiography Certification Exam, Acad Radiol. 32 (2) (2025). 597–603. 10.1016/j.acra.2024.08.009.

[17] T. Kondo, M. Okamoto, Y. Kondo, Pilot Study on Using Large Language Models for Educational Resource Development in Japanese Radiological Technologist Exams, Med Sci Educ. 35 (2) (2025). 919–927. 10.1007/s40670-024-02251-1.

[18] OpenAI, GPT-5 System Card, 2025.

[19] OpenAI, Introducing GPT-5. https://openai.com/index/introducing-gpt-5/, 2025.

[20] S. Wang, M. Hu, Q. Li, M. Safari, X. Yang, Capabilities of GPT-5 on Multimodal Medical Reasoning, arXiv preprint arXiv:2508.08224 (2025).

