## Supplementary material for "Benchmarking GPT-5 Performance and Repeatability on the Japanese National Examination for Radiological Technologists over the Past Decade (2016–2025)"

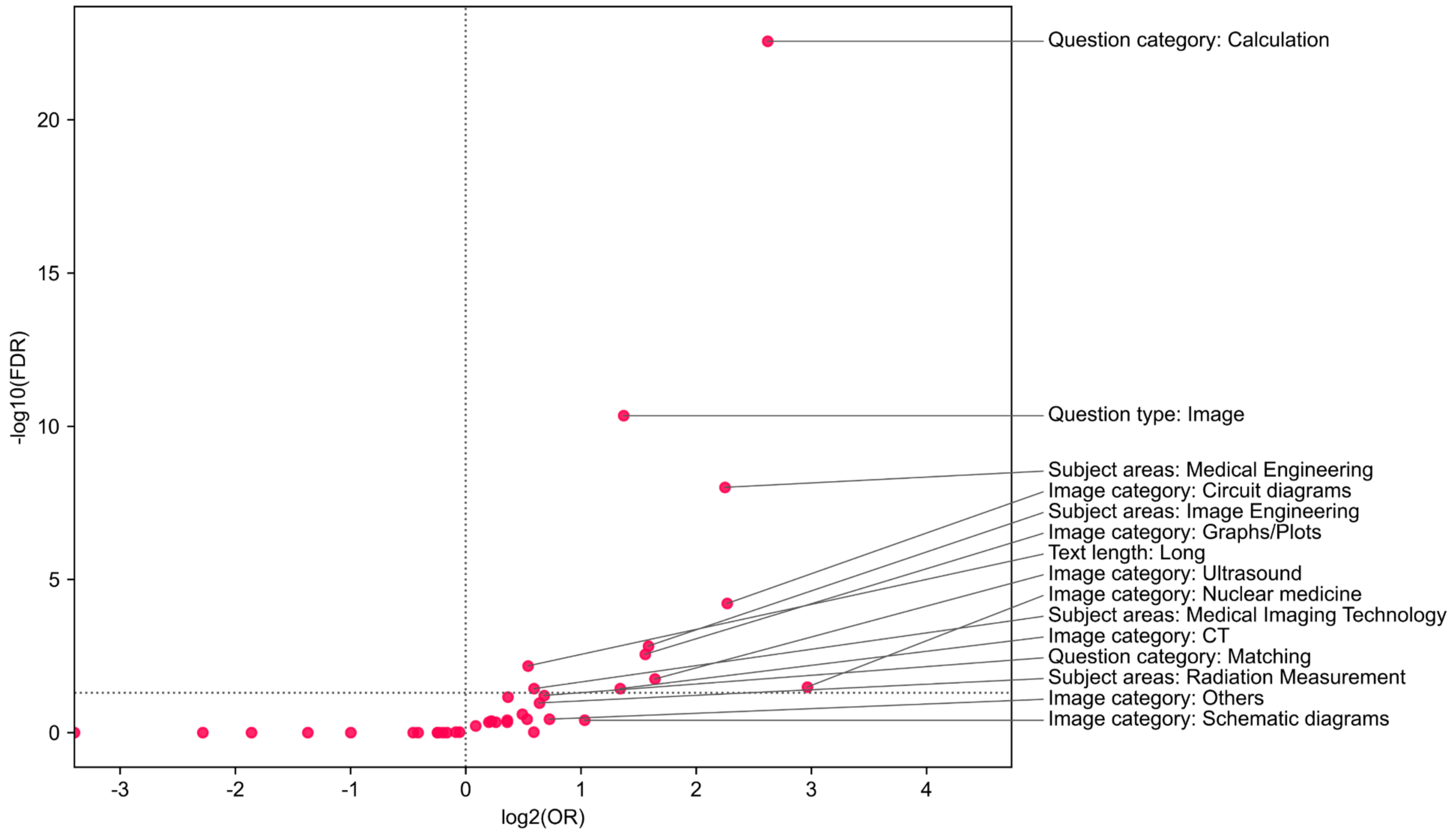


**Supplemental Figure S1.** Volcano plot summarizing the univariable analysis of all candidate factors. The x-axis shows log2 odds ratios (OR), where values greater than zero indicate factors more common among items answered correctly by GPT-5 but incorrectly by GPT-4o. The y-axis shows –log10 false discovery rate–adjusted P values (FDR), with higher values indicating stronger statistical significance. Labeled points represent factors with the largest effect sizes or highest significance.

**Supplemental Table S1.** Repeatability of GPT-5 and GPT-4o across three independent runs on the Japanese national examination for radiological technologists (2016–2025). Results are stratified by question characteristics, subject area, and image category.

| | | GPT-5 | | | | | | | GPT-4o | | | | | | |
|---|---|---|---|---|---|---|---|---|---|---|---|---|---|---|---|
| | | 3-match \| 3/3 correct | 3-match \| 0/3 correct | 2-match \| 2/3 correct | 2-match \| 1/3 correct | 2-match \| 0/3 correct | No-match \| 1/3 correct | No-match \| 0/3 correct | 3-match \| 3/3 correct | 3-match \| 0/3 correct | 2-match \| 2/3 correct | 2-match \| 1/3 correct | 2-match \| 0/3 correct | No-match \| 1/3 correct | No-match \| 0/3 correct |
| Total | Total | 88.2% (1756/1992) | 2.8% (55/1992) | 4.6% (91/1992) | 2.2% (43/1992) | 1.5% (29/1992) | 0.7% (13/1992) | 0.3% (5/1992) | 68.9% (1373/1992) | 19.1% (381/1992) | 3.5% (70/1992) | 3.9% (78/1992) | 4.1% (81/1992) | 0.2% (4/1992) | 0.3% (5/1992) |
| Question type | Text | 93.3% (1569/1682) | 1.3% (22/1682) | 3.2% (53/1682) | 1.1% (18/1682) | 0.7% (12/1682) | 0.3% (5/1682) | 0.2% (3/1682) | 75.3% (1266/1682) | 15.5% (261/1682) | 2.8% (47/1682) | 3.4% (58/1682) | 2.6% (43/1682) | 0.2% (3/1682) | 0.2% (4/1682) |
| | Image | 60.3% (187/310) | 10.6% (33/310) | 12.3% (38/310) | 8.1% (25/310) | 5.5% (17/310) | 2.6% (8/310) | 0.6% (2/310) | 34.5% (107/310) | 38.7% (120/310) | 7.4% (23/310) | 6.5% (20/310) | 12.3% (38/310) | 0.3% (1/310) | 0.3% (1/310) |
| Question style | A type | 88.4% (1499/1696) | 2.7% (46/1696) | 4.8% (81/1696) | 1.9% (33/1696) | 1.4% (23/1696) | 0.6% (11/1696) | 0.2% (3/1696) | 69.0% (1170/1696) | 19.0% (322/1696) | 3.9% (66/1696) | 4.0% (68/1696) | 3.7% (63/1696) | 0.2% (3/1696) | 0.2% (4/1696) |
| | X2 type | 86.8% (257/296) | 3.0% (9/296) | 3.4% (10/296) | 3.4% (10/296) | 2.0% (6/296) | 0.7% (2/296) | 0.7% (2/296) | 68.6% (203/296) | 19.9% (59/296) | 1.4% (4/296) | 3.4% (10/296) | 6.1% (18/296) | 0.3% (1/296) | 0.3% (1/296) |
| Question category | General | 88.4% (1509/1707) | 3.0% (51/1707) | 4.5% (77/1707) | 2.0% (34/1707) | 1.2% (21/1707) | 0.6% (10/1707) | 0.3% (5/1707) | 73.2% (1250/1707) | 17.1% (292/1707) | 3.1% (53/1707) | 3.5% (60/1707) | 2.7% (46/1707) | 0.2% (3/1707) | 0.2% (3/1707) |
| | Calculation | 94.0% (141/150) | 0% (0/150) | 3.3% (5/150) | 0.7% (1/150) | 0.7% (1/150) | 1.3% (2/150) | 0% (0/150) | 30.7% (46/150) | 38.0% (57/150) | 8.7% (13/150) | 6.7% (10/150) | 14.0% (21/150) | 0.7% (1/150) | 1.3% (2/150) |
| | Matching | 78.0% (92/118) | 3.4% (4/118) | 5.9% (7/118) | 6.8% (8/118) | 5.9% (7/118) | 0% (0/118) | 0% (0/118) | 54.2% (64/118) | 25.4% (30/118) | 3.4% (4/118) | 5.9% (7/118) | 11.0% (13/118) | 0% (0/118) | 0% (0/118) |
| | Ordering | 82.4% (14/17) | 0% (0/17) | 11.8% (2/17) | 0% (0/17) | 0% (0/17) | 5.9% (1/17) | 0% (0/17) | 76.5% (13/17) | 11.8% (2/17) | 0% (0/17) | 5.9% (1/17) | 5.9% (1/17) | 0% (0/17) | 0% (0/17) |
| Question wording | Affirmative | 88.2% (1598/1811) | 2.8% (51/1811) | 4.3% (77/1811) | 2.3% (41/1811) | 1.4% (26/1811) | 0.7% (13/1811) | 0.3% (5/1811) | 68.2% (1236/1811) | 19.4% (351/1811) | 3.8% (68/1811) | 3.9% (70/1811) | 4.3% (77/1811) | 0.2% (4/1811) | 0.3% (5/1811) |
| | Negative | 87.3% (158/181) | 2.2% (4/181) | 7.7% (14/181) | 1.1% (2/181) | 1.7% (3/181) | 0% (0/181) | 0% (0/181) | 75.7% (137/181) | 16.6% (30/181) | 1.1% (2/181) | 4.4% (8/181) | 2.2% (4/181) | 0% (0/181) | 0% (0/181) |
| Subject area | Basic Medical Sciences | 98.3% (295/30) | 0.3% (1/30) | 0.7% (2/30) | 0.7% (2/30) | 0% (0/30) | 0% (0/30) | 0% (0/30) | 95.0% (285/30) | 2.7% (8/30) | 1.0% (3/30) | 1.0% (3/30) | 0.3% (1/30) | 0% (0/30) | 0% (0/30) |
| | Diagnostic Imaging | 78.0% (156/20) | 6.5% (13/20) | 6.0% (12/20) | 5.5% (11/20) | 3.0% (6/20) | 1.0% (2/20) | 0% (0/20) | 61.5% (123/20) | 22.0% (44/20) | 3.5% (7/20) | 7.5% (15/20) | 5.0% (10/20) | 0% (0/20) | 0.5% (1/20) |
| | Medical Imaging Technology | 84.0% (168/20) | 2.5% (5/20) | 8.5% (17/20) | 1.5% (3/20) | 3.5% (7/20) | 0% (0/20) | 0% (0/20) | 62.5% (125/20) | 24.0% (48/20) | 2.0% (4/20) | 7.0% (14/20) | 4.0% (8/20) | 0% (0/20) | 0.5% (1/20) |
| | X-ray Imaging Technology | 71.5% (143/20) | 7.0% (14/20) | 10.5% (21/20) | 3.0% (6/20) | 3.5% (7/20) | 3.0% (6/20) | 1.5% (3/20) | 56.0% (112/20) | 27.0% (54/20) | 4.5% (9/20) | 3.5% (7/20) | 8.5% (17/20) | 0.5% (1/20) | 0% (0/20) |
| | Radiation Therapy Technology | 92.4% (182/197) | 2.0% (4/197) | 2.0% (4/197) | 2.0% (4/197) | 0.5% (1/197) | 0.5% (1/197) | 0.5% (1/197) | 71.6% (141/197) | 18.3% (36/197) | 4.1% (8/197) | 3.0% (6/197) | 3.0% (6/197) | 0% (0/197) | 0% (0/197) |
| | Nuclear Medicine Technology | 94.9% (185/195) | 1.5% (3/195) | 1.5% (3/195) | 0.5% (1/195) | 1.0% (2/195) | 0.5% (1/195) | 0% (0/195) | 75.9% (148/195) | 16.4% (32/195) | 3.1% (6/195) | 3.6% (7/195) | 0.5% (1/195) | 0% (0/195) | 0.5% (1/195) |
| | Medical Imaging Informatics | 94.0% (94/10) | 2.0% (2/10) | 1.0% (1/10) | 2.0% (2/10) | 1.0% (1/10) | 0% (0/10) | 0% (0/10) | 67.0% (67/10) | 19.0% (19/10) | 4.0% (4/10) | 6.0% (6/10) | 3.0% (3/10) | 1.0% (1/10) | 0% (0/10) |
| | Radiation Physics | 94.0% (94/10) | 0% (0/10) | 3.0% (3/10) | 3.0% (3/10) | 0% (0/10) | 0% (0/10) | 0% (0/10) | 69.0% (69/10) | 21.0% (21/10) | 4.0% (4/10) | 3.0% (3/10) | 2.0% (2/10) | 0% (0/10) | 1.0% (1/10) |
| | Radiation Measurement | 90% (90/10) | 3.0% (3/10) | 3.0% (3/10) | 4.0% (4/10) | 0% (0/10) | 0% (0/10) | 0% (0/10) | 63.0% (63/10) | 25.0% (25/10) | 2.0% (2/10) | 3.0% (3/10) | 7.0% (7/10) | 0% (0/10) | 0% (0/10) |
| | Radiation Biology | 96.0% (95/99) | 1.0% (1/99) | 3.0% (3/99) | 0% (0/99) | 0% (0/99) | 0% (0/99) | 0% (0/99) | 77.8% (77/99) | 17.2% (17/99) | 2.0% (2/99) | 1.0% (1/99) | 1.0% (1/99) | 1.0% (1/99) | 0% (0/99) |
| | Radiation Safety Management | 74.7% (74/99) | 5.1% (5/99) | 10.1% (10/99) | 5.1% (5/99) | 3.0% (3/99) | 1.0% (1/99) | 1.0% (1/99) | 55.6% (55/99) | 26.3% (26/99) | 5.1% (5/99) | 3.0% (3/99) | 9.1% (9/99) | 1.0% (1/99) | 0% (0/99) |
| | Radiochemistry | 92.0% (69/75) | 1.3% (1/75) | 4.0% (3/75) | 1.3% (1/75) | 0% (0/75) | 1.3% (1/75) | 0% (0/75) | 78.7% (59/75) | 14.7% (11/75) | 2.7% (2/75) | 2.7% (2/75) | 1.3% (1/75) | 0% (0/75) | 0% (0/75) |

| | | | | | | | | | | | | | | | |
|---|---|---|---|---|---|---|---|---|---|---|---|---|---|---|---|
| | Medical Engineering | 89.7% (61/68) | 1.5% (1/68) | 5.9% (4/68) | 1.5% (1/68) | 1.5% (1/68) | 0% (0/68) | 0% (0/68) | 30.9% (21/68) | 35.3% (24/68) | 11.8% (8/68) | 8.8% (6/68) | 13.2% (9/68) | 0% (0/68) | 0% (0/68) |
| | Image Engineering | 82.4% (42/51) | 3.9% (2/51) | 9.8% (5/51) | 0% (0/51) | 2.0% (1/51) | 2.0% (1/51) | 0% (0/51) | 39.2% (20/51) | 31.4% (16/51) | 11.8% (6/51) | 3.9% (2/51) | 11.8% (6/51) | 0% (0/51) | 2.0% (1/51) |
| | Patient Safety Management | 10.0% (8/8) | 0% (0/8) | 0% (0/8) | 0% (0/8) | 0% (0/8) | 0% (0/8) | 0% (0/8) | 10.0% (8/8) | 0% (0/8) | 0% (0/8) | 0% (0/8) | 0% (0/8) | 0% (0/8) | 0% (0/8) |
| Image category | MRI | 59.0% (36/61) | 16.4% (10/61) | 8.2% (5/61) | 9.8% (6/61) | 4.9% (3/61) | 1.6% (1/61) | 0% (0/61) | 49.2% (30/61) | 37.7% (23/61) | 3.3% (2/61) | 3.3% (2/61) | 6.6% (4/61) | 0% (0/61) | 0% (0/61) |
| | X-ray | 51.8% (29/56) | 17.9% (10/56) | 12.5% (7/56) | 3.6% (2/56) | 5.4% (3/56) | 5.4% (3/56) | 3.6% (2/56) | 33.9% (19/56) | 41.1% (23/56) | 7.1% (4/56) | 7.1% (4/56) | 10.7% (6/56) | 0% (0/56) | 0% (0/56) |
| | CT | 47.1% (16/34) | 17.6% (6/34) | 23.5% (8/34) | 2.9% (1/34) | 5.9% (2/34) | 2.9% (1/34) | 0% (0/34) | 32.4% (11/34) | 32.4% (11/34) | 5.9% (2/34) | 8.8% (3/34) | 20.6% (7/34) | 0% (0/34) | 0% (0/34) |
| | Ultrasound | 46.4% (13/28) | 10.7% (3/28) | 17.9% (5/28) | 10.7% (3/28) | 10.7% (3/28) | 3.6% (1/28) | 0% (0/28) | 21.4% (6/28) | 35.7% (10/28) | 10.7% (3/28) | 14.3% (4/28) | 14.3% (4/28) | 0% (0/28) | 3.6% (1/28) |
| | Nuclear medicine | 85.7% (6/7) | 0% (0/7) | 0% (0/7) | 0% (0/7) | 14.3% (1/7) | 0% (0/7) | 0% (0/7) | 0% (0/7) | 57.1% (4/7) | 14.3% (1/7) | 28.6% (2/7) | 0% (0/7) | 0% (0/7) | 0% (0/7) |
| | Fundus | 75.0% (3/4) | 0% (0/4) | 0% (0/4) | 25.0% (1/4) | 0% (0/4) | 0% (0/4) | 0% (0/4) | 50% (2/4) | 50% (2/4) | 0% (0/4) | 0% (0/4) | 0% (0/4) | 0% (0/4) | 0% (0/4) |
| | Graphs/Plots | 59.6% (28/47) | 6.4% (3/47) | 19.1% (9/47) | 12.8% (6/47) | 2.1% (1/47) | 0% (0/47) | 0% (0/47) | 25.5% (12/47) | 46.8% (22/47) | 12.8% (6/47) | 2.1% (1/47) | 10.6% (5/47) | 2.1% (1/47) | 0% (0/47) |
| | Circuit diagrams | 88.6% (31/35) | 2.9% (1/35) | 5.7% (2/35) | 2.9% (1/35) | 0% (0/35) | 0% (0/35) | 0% (0/35) | 31.4% (11/35) | 31.4% (11/35) | 11.4% (4/35) | 5.7% (2/35) | 20% (7/35) | 0% (0/35) | 0% (0/35) |
| | Schematic diagrams | 64.3% (9/14) | 0% (0/14) | 0% (0/14) | 28.6% (4/14) | 7.1% (1/14) | 0% (0/14) | 0% (0/14) | 35.7% (5/14) | 28.6% (4/14) | 0% (0/14) | 14.3% (2/14) | 21.4% (3/14) | 0% (0/14) | 0% (0/14) |
| | Others | 65.6% (21/32) | 3.1% (1/32) | 9.4% (3/32) | 6.2% (2/32) | 9.4% (3/32) | 6.2% (2/32) | 0% (0/32) | 43.8% (14/32) | 34.4% (11/32) | 9.4% (3/32) | 3.1% (1/32) | 9.4% (3/32) | 0% (0/32) | 0% (0/32) |

3-match | 3/3 correct = all three runs were identical and correct, 3-match | 0/3 correct = all three runs were identical and incorrect, 2-match | 2/3 correct = two of the three runs were identical and correct, 2-match | 1/3 correct = two of the three runs were identical and incorrect, 2-match | 0/3 correct = two of the three runs were identical and incorrect, while the third was also incorrect but a different wrong option, No-match | 1/3 correct = all three runs produced different outputs, with one correct, No-match | 0/3 correct = all three runs produced different outputs, none correct.
